# Role of age-friendly communities on the association between informal caregiving and depressive symptoms: a multilevel analysis using the Japan Gerontological Evaluation Study

**DOI:** 10.1101/2025.05.23.25328052

**Authors:** Taiji Noguchi, Satoko Fujihara, Kazushige Ide, Seungwon Jeong, Tami Saito, Katsunori Kondo, Toshiyuki Ojima

**Affiliations:** Department of Community Health and Preventive Medicine, Hamamatsu University School of Medicine, Hamamatsu, Japan; Department of Social Science, Research Institute, National Center for Geriatrics and Gerontology, Aichi, Japan; Institute for Health Economics and Policy, Association for Health Economics Research and Social Insurance and Welfare, Tokyo, Japan; Center for Preventive Medical Sciences, Chiba University, Chiba, Japan; Research Team for Promoting Independence and Mental Health, Tokyo Metropolitan Institute for Geriatrics and Gerontology, Tokyo, Japan; Department of Community Building for Well-being, Center for Preventive Medical Sciences, Chiba University, Chiba, Japan; Department of Community Welfare, Niimi University, Okayama, Japan

**Keywords:** Age-friendliness, Dementia-friendly communities, Carer, Unpaid caregiving, Mental health

## Abstract

**Background and Objectives:** Age-friendly communities (AFC) are inclusive and accessible community environments that ensure older adults’ quality of life and dignity. However, empirical evidence regarding the AFC influence on informal caregivers is lacking. This study examined the moderating role of AFC on the association between caregiving and depressive symptoms.

**Research Design and Methods:** This cross-sectional study included 10,315 older adults (mean 74.2 years) across 145 communities from the Japan Gerontological Evaluation Study 2016 data. Participants were divided based on providing care to families: “not caregivers” and “caregivers.” The AFC scale assessed community-level age-friendliness in three domains: age-friendly physical environments (accessibility to barrier-free outdoor spaces, buildings, and transportation), social engagement and communication (participation in community groups, volunteer engagement, and information use), and social inclusion and dementia- friendliness (respect and inclusion for older adults and people with dementia). The multilevel analysis with individuals nested in communities examined the cross-level interactions of caregiving and community-level age-friendliness on depressive symptoms assessed using the Geriatrics Depression Scale-15.

**Results:** Among the participants, 7.1% were caregivers. Caregiving was associated with higher depressive symptoms. Of the AFC scale, the social inclusion and dementia-friendliness domain moderated the association between caregiving and depressive symptoms, whereas the social engagement and communication domain amplified this association. The age-friendly physical environment domain showed no modifications.

**Discussion and Implications:** Inclusive and dementia-friendly aspects of AFC potentially moderate the caregivers’ depressive symptoms. However, the community’s physical environment and social interaction elements may not relate to caregiver’s mental benefits. These findings underscore the AFC promotion considering caregivers.

## Introduction

Informal caregivers are anyone who provides help and care, without remuneration, to family members or friends due to illness, disability, addiction, or mental health problems (Carers Trust, n.d.). They play a major role in increasing essential care resources worldwide. The United Nations estimates that more than 70% of care needs are met by informal caregivers (United Nations Economic Commission for Europe, 2019). In the United States, approximately 41.8 million people provided unpaid care to persons aged 50 and older in 2020 (AARP and National Alliance for Caregiving, 2020). Japan, a super-aged society, has approximately 6.5 million family caregivers for adults under long-term care (Statistics Bureau of Japan, 2022). With the population aging, the need for informal caregivers is expected to increase. Therefore, developing support measures for them is a crucial international public health and gerontological challenge.

Informal caregivers experience several care-related burdens, including adverse influences on their emotional, social, financial, physical, and spiritual functioning (Adelman et al., 2014; Zarit et al., 1986). Particularly, mental health problems are a major concern to caregivers (Ervin et al., 2022), who are reported as more likely to be affected by caregiver burden than physical and financial impacts (Yates et al., 1999). Caregivers suffer from approximately 30–50% of mental health problems, such as depressive symptoms, anxiety, and suicidal ideation (Collins & Kishita, 2020; Mohamad et al., 2024; Yao et al., 2024). Theoretically, several care-related models suggest that caregivers’ mental health decline is caused by an interaction between stressors (e.g., care recipients’ functional conditions, such as cognitive impairment), appraisals for the stressors (e.g., caregiving time or overload), and resources (e.g., formal and informal support) (Pearlin et al., 1990; Verbakel et al., 2018; Yates, et al., 1999). To protect caregivers’ mental health, it is important to explore resources which moderate these stressors and burdens as well as identify the stressors and their causes.

Age-friendly communities (AFC), promoted by the World Health Organization (WHO) to achieve healthy aging, is an international initiative that reflects the commitment to inclusive and accessible community environments (WHO, 2015). The AFC optimizes opportunities for health, participation, and safety for all individuals, ensuring the quality of life and dignity as people age (Meeks, 2022; WHO, 2015). Given that caregivers and their care recipients are one of the groups that may require greater resources and support from the community (Black et al., 2016), the nature of the community’s age-friendliness can work as a moderating role in the stressors and burdens experienced by caregivers. The AFC consists of eight topic areas that comprehensively cover the age-friendliness of communities, including the features of the structures, environments, services, and policies of communities that reflect the determinants of healthy aging (WHO, 2007, 2015, 2020). It includes built environments, such as outdoor spaces and transportation, access to supportive community health services, availability of social participation, and social environments, such as the inclusion and respect of older adults.

Caregiving is an integrative relationship that incorporates not only the caregiver, the care recipient, and the family unit, but also the surrounding community as well. Thus, it is crucial to include the relationship between the caregiver and the context of their community environment to consider caregivers’ personal needs and well-being. In light of the theory of care-related models, AFC environments may serve as a resource to buffer responses to stressors. Providing support to caregivers and including them in the community’s social and physical environments may alleviate mental health programs by decreasing caregiver stress and burden and increasing community participation and inclusion (Papastavrou et al., 2015; Williams et al., 2017). Several previous studies have suggested that community environments and neighborhood characteristics help caregivers’ health and well-being. Higher levels of community social capital (i.e., social environmental factors such as residents’ trust and participation) moderated caregivers’ mental health decline (Mohanty et al., 2020). The rich social relationships in the neighborhood suppressed psychological distress due to the severe caregiving burden (Noguchi et al., 2020a). Meanwhile, caregivers and their care recipients often report built environments, such as transportation, as barriers to healthcare, causing undue distress (Black, et al., 2016). These support the potential of environmental features such as AFC to help caregivers for mental health issues.

However, despite that WHO initially included caregiver-friendly in the AFC (WHO, 2007), little is known to evaluate the AFC impact on caregivers. Communities with inclusiveness and accessibility may help caregivers obtain social support and assist the care recipients’ travel, and reduce the burden related to care (Mohanty et al., 2020). This perspective suggests that AFC may benefit especially vulnerable populations such as caregivers, rather than the older population in general. Nevertheless, previous research has been limited to surveys of caregivers’ perception of AFC, such as their satisfaction with AFC (Black et al., 2016; Johnson et al., 2016), and there is a lack of empirical evidence of the influence of AFC on informal caregivers (Liebzeit et al., 2023). Considering that developing communities with caregiver-friendly may also contribute to the quality of life and well-being of older adults with functional limitations and dementia, it is useful for public health and urban planning policymakers to understand the impact of AFC on caregivers and to capture which aspects of AFC would benefit caregivers.

In this light, this study aimed to examine the moderating role of AFC on the association between informal caregiving and depressive symptoms. Specifically, we used a validated AFC scale to assess community-level age-friendliness and explored the moderation of community-level factors through a multilevel analysis.

## Methods

### Data

This cross-sectional study used data from the Japan Gerontological Evaluation Study (JAGES) 2016 wave, which is an ongoing cohort study for older adults aged 65 years and above in Japan, who were not certified as needing long-term care (Kondo et al., 2018). In Japan, a public long-term care insurance system was launched in 2000, and older adults aged 65 and above (or those aged 40–64 who require long-term care due to certain diseases, such as terminal cancer, Alzheimer’s disease, and stroke) are eligible to receive long-term care services based on their needs throughout the county (Konishi et al., 2024).

Figure 1 illustrates the flow of sample selection. Between October and December 2016, self-administered questionnaires were distributed through mail to older adults who lived in 39 municipalities. The municipalities were not randomly selected but covered a wide range of regions and population sizes in Japan. Among them, random sampling methods were used in 17 large municipalities, while complete sampling methods were used in 22 other municipalities. Of the participants, several items related to the AFC scale were randomly assigned to the survey questionnaire modules in one-eighth. Consequently, a total of 270,661 individuals were invited to participate in the survey; 196,438 returned the questionnaires (response rate = 70.2%), among which, 16,417 respondents with invalid information on age and/or sex or who received public long-term care insurance benefits were excluded (valid response rate = 64.4%). We defined the school districts as community units because school districts in Japan are reasonable units for considering local public health activities, as older adults can easily travel on foot or by bicycle (Takagi et al., 2013). In order to avoid imprecision due to the small sample size, we excluded 87,511 individuals across 849 community areas with less than 30 respondents (Tsuji et al., 2018), because AFC-related survey modules assigned part of the participants, and 9,899 respondents with unknown areas of residence were also excluded. Therefore, data from 82,611 participants across 145 communities were used to calculate AFC scale scores at the community level. Furthermore, to identify informal caregivers, we restricted the participants to those assigned to the survey module on caregiving (which was randomly assigned); thus, 72,296 individuals who were not assigned to that survey module were excluded. Consequently, a total of 10,315 individuals across 145 communities were included in the final analysis.

**Figure 1.**
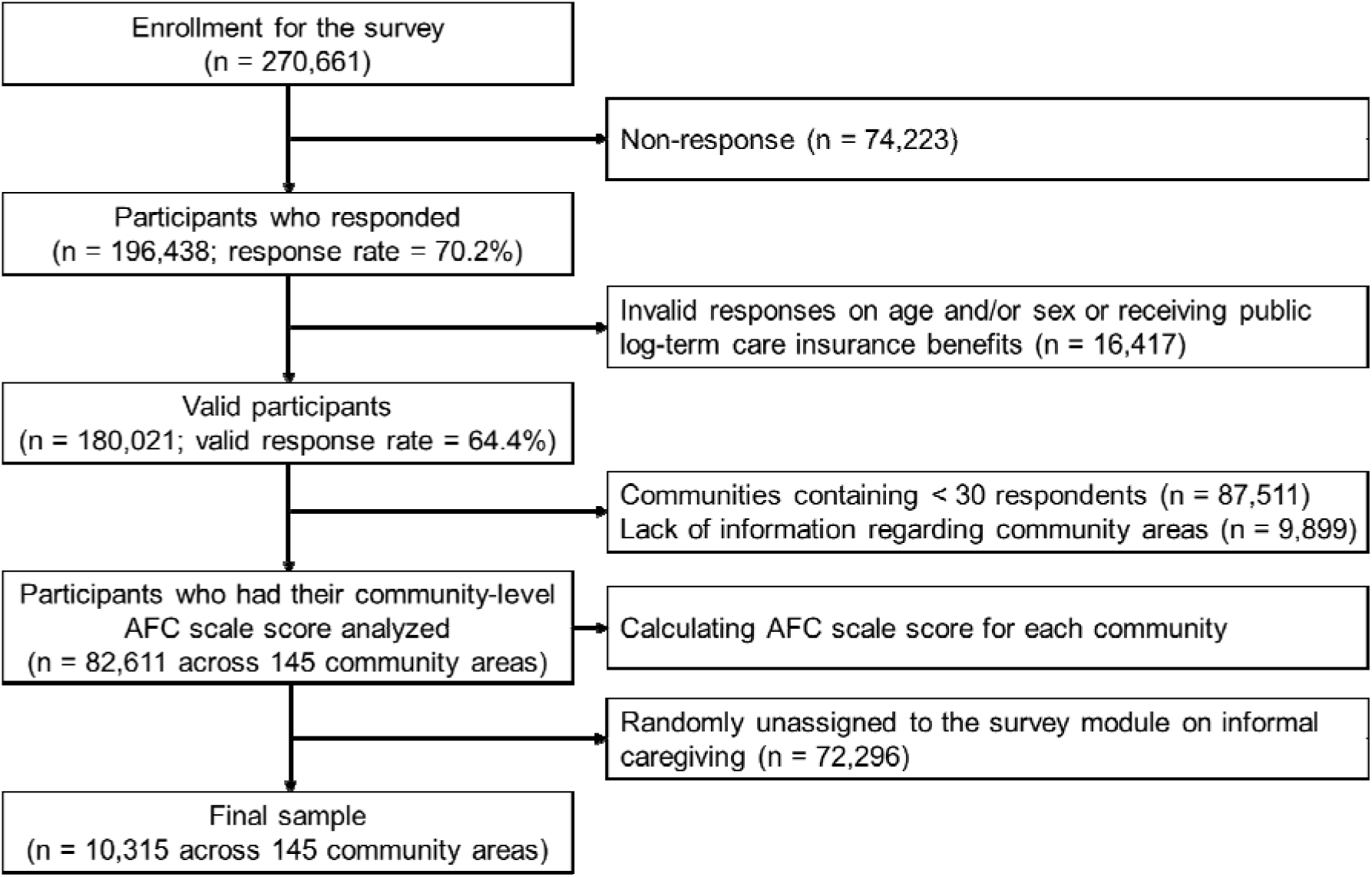
Sample selection flow chart. AFC, age-friendly community

This study was reviewed and approved by the Ethics committees at the National Center for Geriatrics and Gerontology (No. 992) and Chiba University (No. 2493). The mailed questionnaire was accompanied by a study explanation, and the participants who returned the completed questionnaire were considered to have provided informed consent. All the procedures were performed in accordance with the Declaration of Helsinki.

### Depressive symptoms

Depressive symptoms were measured using the Japanese version of the Geriatric Depression Scale, 15-point edition (GDS-15) (D’Ath et al., 1994; Wada et al., 2004). This measure was developed to assess depressive symptoms in self-administered surveys. This scale consists of 15 items with yes and no answer options that yield a total score ranging from 0 to 15, with higher scores indicating more depressive symptoms. In this study, we used this score as a continuous measure.

### Caregiving status

Caregiving status was assessed using the question, “Do you currently provide care for your family?” and the participants were divided into two groups: “not caregivers” and “caregivers.”

Subsequently, we further divided for the secondary consideration of caregivers in terms of caregiving for persons with dementia and caregiving role. Regarding care for persons with dementia, caregivers were asked an additional question, “Does the person you care for have symptoms of dementia?” with possible responses of “no,” “yes (symptomatic but not yet diagnosed)”, and “yes (diagnosed).” Based on the answer, the caregivers were further divided to “caregivers for persons without dementia” and “caregivers for persons with dementia” (symptomatic but not yet diagnosed or diagnosed). Regarding the caregiving role, the caregivers were further divided into “secondary caregiver” (not mainly providing care, but supporting the primary caregiver in relation to care) and “primary caregiver” (mainly providing care).

### Community-level age-friendliness

Based on previous studies (Fujihara et al., 2025; Noguchi et al., 2024), we measured community age-friendliness, using the AFC scale which comprises three domains: (i) age-friendly physical environments, (ii) social engagement and communication, and (iii) social inclusion and dementia-friendliness. This scale encompasses the AFC elements proposed by the WHO and incorporates dementia-friendly aspects, and its validity and reliability have been certified (Fujihara et al., 2025). We calculated age-friendliness scores for each community by aggregating individual responses on the AFC scale items by school districts which are used as community units (Takagi et al., 2013). Supplementary Table 1 presents the details of the scale items. The domain of age-friendly physical environments evaluated accessibility to barrier-free outdoor spaces and buildings and transportation (five items: score range 0–500 points). The domain of social engagement and communication assessed the level of older adults’ social participation and volunteer engagement in the community and the abundance of information communication (five items: score range 0–500 points). Finally, the domain of social inclusion and dementia-friendliness measured the elements of respect and inclusion for older adults and dementia-friendly beliefs and attitudes (seven items: score range 0–700 points). The higher scores for each domain indicate more age-friendly.

### Covariates

The covariates included age, gender, living arrangement, marital status, educational attainment, equivalent household income, home ownership, total household assets, working frequency, number of illnesses, basic and instrumental activities of daily living (BADL and IADL) performance, and community-level population density, aging proportion, low education proportion, and care resources. Age (years) was categorized as 65–69, 70–74, 75– 79, 80–84, and ≥ 85. Living arrangement was dichotomized as living with others and living alone. Marital status was categorized as married, widowed/divorced, never married, and other. Educational attainment (years) was categorized as < 10, 10–12, and ≥ 13. Equivalent household income was calculated by dividing the income of each household by the square root of the number of family members, and categorized as low (< 2.00 million JPY), middle (2.00–3.00 million JPY), and high (≥ 3.00 million JPY). Home ownership was dichotomized as yes or no. Total household assets (including savings, real estate, and stocks) were divided into three groups: low (< 10 million JPY), middle (10–50 million JPY), and high (≥ 50 million JPY). Working frequency was categorized as not working, sometimes, weekly, and almost every day. The number of illnesses was categorized as none, one, two, and three or more, from a list of 17 illnesses, such as cancer, heart disease, stroke, diabetes, digestive diseases, and musculoskeletal disorders. BADL performance was assessed by the question, “Do you need care or assistance in your daily life?” and the participants were divided into those without difficulty (not requiring care or assistance) and those with difficulty (requiring some kind of care or assistance). IADL performance was assessed using a five-item subscale of the Tokyo Metropolitan Institute of Gerontology Index of Competence (Koyano et al., 1991); those with at least one item of difficulty were defined as with difficulty while others were without difficulty.

The community-level covariates included population density (divided into quartiles), aging proportion (the proportion of people aged ≥ 65 years; divided into quartiles), and low education proportion (the proportion of people who had graduated only from junior high school; divided into quartiles); the information was based on municipal-level data from the Japanese Census data. Care resource levels in the community were also included as the covariates (Takahashi et al., 2015). We calculated the total number of care service providers in each municipality for home-visit care services, outpatient care services, short-stay care services, and home care support offices (care plan centers), and defined the care resource level of care service providers for every 1,000 older adults in that community (divided into quartiles), which was applied as a community-level variable.

### Statistical analysis

First, the descriptive statistics of the characteristics of the individuals and communities were summarized. Second, we conducted a multilevel linear regression analysis with a hierarchical structure of the first level as individuals and the second level as communities and obtained regression coefficients and 95% confidence intervals (CIs) for depressive symptom scores. The multilevel framework assumes that individual outcomes are partially dependent on the communities in which the individuals live. Initially, we used a null model to assess whether depressive symptom scores varied across communities. Next, we examined the association between caregiving status and depressive symptoms, adjusted for the covariates, including community-level variables. Subsequently, we included a cross-level interaction term of caregiving status and community-level AFC scale scores in the analytical model to explore the moderation of AFC on the association between caregiving status and depressive symptoms. In the analysis, the AFC score was centralized, and the estimates were made for every ten percent point of the AFC score. Finally, the predicted value of depressive symptoms for each AFC scale domain was plotted according to caregiving status.

Additionally, we performed a subgroup analysis by gender. Furthermore, we exploratively conducted a similar analytical model for additional divided caregiving statuses (caregiving for persons with dementia and caregiving role).

To mitigate potential bias due to missing information, we performed missing-value imputation using chained random forests, based on the random forest algorithm (Stekhoven & Bühlmann, 2012).

The significance level was set at < 0.05. We used the R software (Version 4.3.1 for Windows; R Foundation for Statistical Computing, Vienna, Austria) for all statistical analyses.

## Results

Data from 10,315 individuals across 145 communities were analyzed. Table 1 presents the characteristics of the participants. The mean age was 74.2 years (standard deviation [SD] = 6.4), and 54.4% were females. Of the participants, 733 (7.1%) were engaged in informal caregiving. The caregivers were more likely to be female, live with others, married, and have less difficulty in IADL performance. Among the caregivers, 54.6% provided care to families with dementia, and 49.8% were primarily engaged in caregiving. The caregivers had higher depressive symptom scores than non-caregivers (mean [SD]: caregivers = 3.2 [3.2]; not caregivers = 2.8 [3.0]).

**Table 1.** Characteristics of the participants.

|  |  | Overall | Caregiving status* |  |
| --- | --- | --- | --- | --- |
|  |  |  | Not caregivers | Caregivers |
|  |  | n = 10315 | n = 8362 | n = 733 |
| Age (years), n (%) | 65 to 69 | 3193 (31.0) | 2683 ( 32.1) | 303 ( 41.3) |
|  | 70 to 74 | 2632 (25.5) | 2264 ( 27.1) | 144 ( 19.6) |
|  | 75 to 79 | 2263 (21.9) | 1834 ( 21.9) | 131 ( 17.9) |
|  | 80 to 84 | 1426 (13.8) | 1039 ( 12.4) | 108 ( 14.7) |
|  | 85 and older | 801 ( 7.8) | 542 ( 6.5) | 47 ( 6.4) |
| Gender, n (%) | Men | 4706 (45.6) | 3944 ( 47.2) | 289 ( 39.4) |
|  | Women | 5609 (54.4) | 4418 ( 52.8) | 444 ( 60.6) |
| Living arrangement, n (%) | Living with others | 8337 (80.8) | 6908 ( 82.6) | 642 ( 87.6) |
|  | Living alone | 1305 (12.7) | 1010 ( 12.1) | 36 ( 4.9) |
|  | Missing | 673 ( 6.5) | 444 ( 5.3) | 55 ( 7.5) |
| Marital status, n (%) | Married | 7384 (71.6) | 6085 ( 72.8) | 635 ( 86.6) |
|  | Separated/divorced | 2430 (23.6) | 1956 ( 23.4) | 66 ( 9.0) |
|  | Never married | 215 ( 2.1) | 173 ( 2.1) | 14 ( 1.9) |
|  | Other | 83 ( 0.8) | 54 ( 0.6) | 3 ( 0.4) |
|  | Missing | 203 ( 2.0) | 94 ( 1.1) | 15 ( 2.0) |
| Educational attainment (years), n (%) | < 10 | 4098 (39.7) | 3162 ( 37.8) | 270 ( 36.8) |
|  | 10 to 12 | 3988 (38.7) | 3359 ( 40.2) | 297 ( 40.5) |
|  | ≥ 13 | 2099 (20.3) | 1774 ( 21.2) | 158 ( 21.6) |
|  | Missing | 130 ( 1.3) | 67 ( 0.8) | 8 ( 1.1) |
| Equivalent household income, n (%) | Low | 3895 (37.8) | 3179 ( 38.0) | 297 ( 40.5) |
|  | Middle | 2881 (27.9) | 2512 ( 30.0) | 206 ( 28.1) |
|  | High | 804 ( 7.8) | 703 ( 8.4) | 56 ( 7.6) |
|  | Missing | 2735 (26.5) | 1968 ( 23.5) | 174 ( 23.7) |
| Household total assets, n (%) | Low | 1910 (18.5) | 1551 ( 18.5) | 138 ( 18.8) |
|  | Middle | 4532 (43.9) | 3878 ( 46.4) | 338 ( 46.1) |
|  | High | 976 ( 9.5) | 860 ( 10.3) | 71 ( 9.7) |
|  | Missing | 2897 (28.1) | 2073 ( 24.8) | 186 ( 25.4) |
| Home ownership, n (%) | No | 771 ( 7.5) | 611 ( 7.3) | 46 ( 6.3) |
|  | Yes | 9390 (91.0) | 7671 ( 91.7) | 679 ( 92.6) |
|  | Missing | 154 ( 1.5) | 80 ( 1.0) | 8 ( 1.1) |
| Working frequency, n (%) | Everyday | 1544 (15.0) | 1315 ( 15.7) | 97 ( 13.2) |
|  | Weekly | 818 ( 7.9) | 691 ( 8.3) | 52 ( 7.1) |
|  | Sometimes | 568 ( 5.5) | 475 ( 5.7) | 46 ( 6.3) |
|  | Not working | 6159 (59.7) | 5080 ( 60.8) | 458 ( 62.5) |
|  | Missing | 1226 (11.9) | 801 ( 9.6) | 80 ( 10.9) |
| BADL performance, n (%) | Without difficulty | 8948 (86.7) | 7431 ( 88.9) | 633 ( 86.4) |
|  | With difficulty | 648 ( 6.3) | 445 ( 5.3) | 63 ( 8.6) |
|  | Missing | 719 ( 7.0) | 486 ( 5.8) | 37 ( 5.0) |
| IADL performance, n (%) | Without difficulty | 5298 (51.4) | 4231 ( 50.6) | 475 ( 64.8) |
|  | With difficulty | 4662 (45.2) | 3905 ( 46.7) | 241 ( 32.9) |
|  | Missing | 355 ( 3.4) | 226 ( 2.7) | 17 ( 2.3) |
| Number of illnesses, n (%) | None | 2571 (24.9) | 2115 ( 25.3) | 209 ( 28.5) |
|  | One | 3905 (37.9) | 3140 ( 37.6) | 264 ( 36.0) |
|  | Two | 2223 (21.6) | 1818 ( 21.7) | 155 ( 21.1) |
|  | Three or more | 1146 (11.1) | 963 ( 11.5) | 79 ( 10.8) |
|  | Missing | 470 ( 4.6) | 326 ( 3.9) | 26 ( 3.5) |
| Caregiving for persons with dementia | Caregivers for a person without dementia | 295 ( 2.9) | 0 ( 0.0) | 295 ( 40.2) |
|  | Caregivers for a person with dementia | 400 ( 3.9) | 0 ( 0.0) | 400 ( 54.6) |
|  | Missing | 1258 (12.2) | 0 ( 0.0) | 38 ( 5.2) |
| Caregiving role | Secondary caregivers | 368 ( 3.6) | 0 ( 0.0) | 368 ( 50.2) |
|  | Primary caregivers | 365 ( 3.5) | 0 ( 0.0) | 365 ( 49.8) |
|  | Missing | 1220 (11.8) | 0 ( 0.0) | 0 ( 0.0) |
| Depressive symptoms score, mean (SD)* |  | 2.9 (3.0) | 2.8 (3.0) | 3.2 (3.2) |

*Note:* BADL, basic activities of daily living; IADL, instrumental activities of daily living; SD, standard deviation.
\*Missing data: n = 1220 (caregiving status); n = 1902 (depressive symptoms score).

Table 2 shows the descriptive statistics for the AFC scale and other community-level covariates. The AFC scale scores ranged from 72.5–273.3 (mean [SD] = 147.6 [38.6]) for age-friendly physical environments, 58.8–173.7 (mean [SD] = 118.2 [22.5]) for social engagement and communication, and 306.6–525.7 (mean [SD] = 386.4 [41.2]) for social inclusion and dementia-friendliness.

**Table 2.**
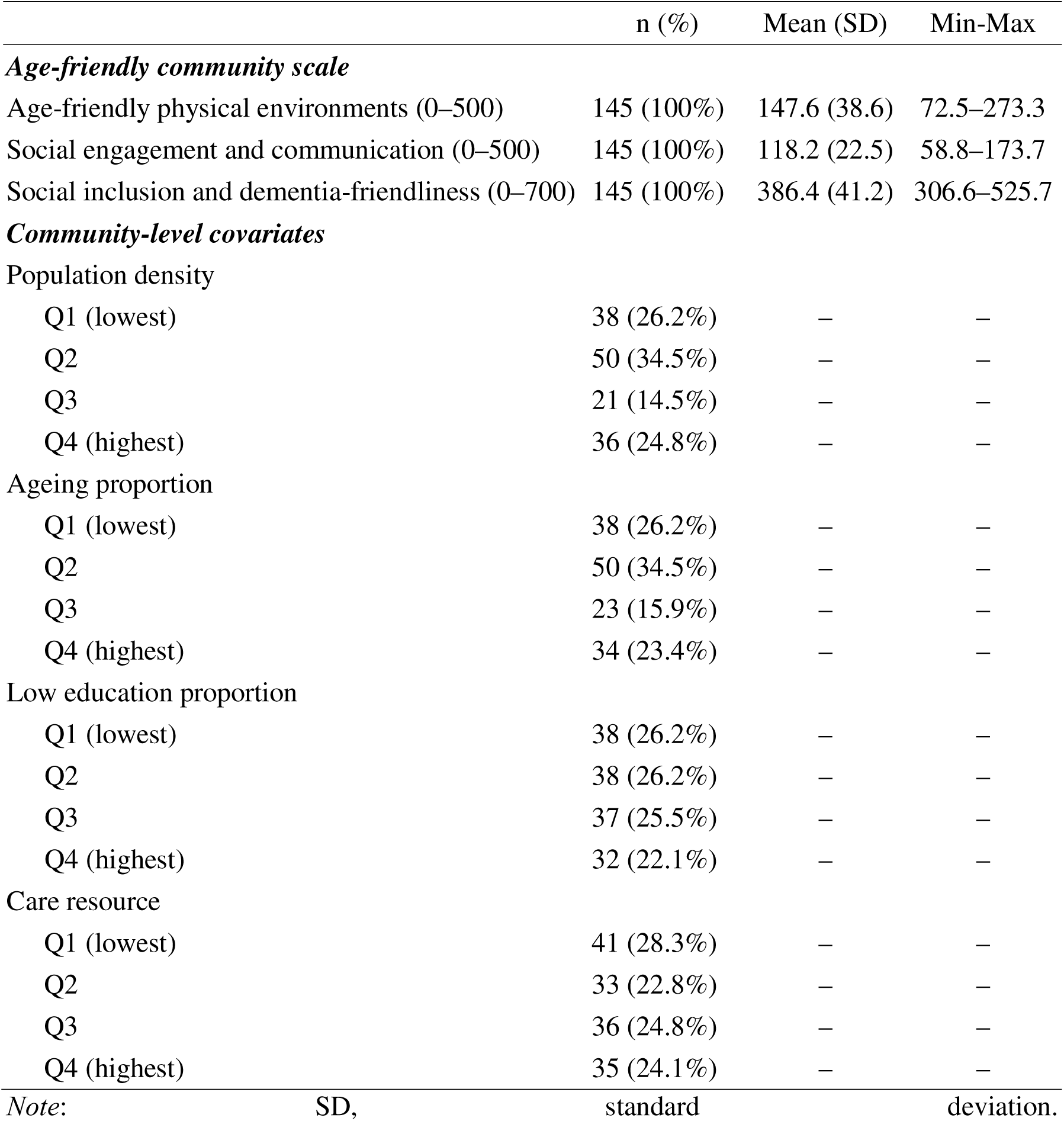
Characteristics of the communities (n = 145)

Table 3 presents the results of the association between caregiving status and depressive symptoms, based on a multilevel linear regression analysis. After adjusting for the covariates, informal caregiving was found to be associated with higher levels of depressive symptoms (Model 2: coef. = 0.36; 95% CI = 0.16, 0.56). Models 3 to 5 indicate the results when including the interaction term of caregiving and AFC scores, and Model 4 showed a positive interaction of caregiving status and social engagement and communication on depressive symptoms, indicating amplification of this association (coef. = 0.59; 95% CI = 0.08, 1.11; *P* for interaction = 0.023). Meanwhile, Model 5 showed a negative interaction between caregiving status and social inclusion and dementia-friendliness on depressive symptoms, with attenuated association (coef. = -0.52; 95% CI = -0.89, -0.15; *P* for interaction = 0.006).

**Table 3.** Association between caregiving status and depressive symptoms, based on multilevel linear regression analysis.

|  | Model 1 |  | Model 2 |  | Model 3 |  | Model 4 |  | Model 5 |  |
| --- | --- | --- | --- | --- | --- | --- | --- | --- | --- | --- |
|  | Coef. (95% CI) | P-value | Coef. (95% CI) | P-value | Coef. (95% CI) | P-value | Coef. (95% CI) | P-value | Coef. (95% CI) | P-value |
| **Fixed effects** |  |  |  |  |  |  |  |  |  |  |
| **Caregiving status** |  |  |  |  |  |  |  |  |  |  |
| Not caregivers | Reference |  | Reference |  | Reference |  | Reference |  | Reference |  |
| Caregivers | 0.34 (0.14, 0.55) | < 0.001 | 0.36 (0.16, 0.56) | < 0.001 | 0.37 (0.17, 0.56) | < 0.001 | 0.37 (0.17, 0.57) | < 0.001 | 0.36 (0.16, 0.56) | < 0.001 |
| **AFC scale** |  |  |  |  |  |  |  |  |  |  |
| Age-friendly physical environments |  |  | 0.00 (-0.11, 0.11) | 0.963 | -0.02 (-0.13, 0.10) | 0.781 | 0.00 (-0.11, 0.11) | 0.938 | 0.00 (-0.11, 0.11) | 0.962 |
| Social engagement and communication |  |  | -0.05 (-0.28, 0.17) | 0.650 | -0.05 (-0.28, 0.17) | 0.644 | -0.11 (-0.34, 0.12) | 0.356 | -0.06 (-0.28, 0.17) | 0.625 |
| Social inclusion and dementia-friendliness |  |  | -0.23 (-0.38, -0.08) | 0.003 | -0.23 (-0.38, -0.08) | 0.003 | -0.23 (-0.38, -0.08) | 0.003 | -0.18 (-0.34, -0.03) | 0.020 |
| **Caregiving status x AFC scale** |  |  |  |  |  |  |  |  |  |  |
| Caregivers x Age-friendly physical environments |  |  |  |  | 0.21 (-0.10, 0.52) | 0.184 |  |  |  |  |
| Caregivers x Social engagement and communication |  |  |  |  |  |  | 0.59 (0.08, 1.11) | 0.023 |  |  |
| Caregivers x Social inclusion and dementia-friendliness |  |  |  |  |  |  |  |  | -0.52 (-0.89, -0.15) | 0.006 |
| **Random effects** |  |  |  |  |  |  |  |  |  |  |
| Community-level variance | 0.08641 |  | 0.04237 |  | 0.04219 |  | 0.04289 |  | 0.04206 |  |

*Note:* AFC, age-friendly community; Coef., unstandardized regression coefficient; CI, confidence interval.
Model 1: crude model; Model 2: adjusted by age, gender, living arrangement, marital status, equivalent household income, home ownership, household total assets, work frequency, number of illnesses, basic and instrumental activities of daily living, population density, aging proportion, low education proportion, care resource, and AFC scale score; Model 3 to 5: added the interaction terms to Model 2 (Model 3: caregiving status x Age-friendly physical environments; Model 4: caregiving status x Social engagement and communication; Model 5: caregiving status x Social inclusion and dementia-friendliness).
Random effects for the null model (community-level variance) = 0.08898.
The AFC scale scores were estimated per ten percent points.
Missing data were imputed by random forest imputation algorithm.

Figure 2 illustrates the predicted value of depressive symptoms for each AFC scale domain according to caregiving status. Regarding the domains of age-friendly physical environments and social engagement and communication, depressive symptoms of non-caregivers decreased, while that decrease in caregivers was not observed. Meanwhile, in communities with higher scores on the domain of social inclusion and dementia-friendliness, caregivers’ depressive symptoms gradually lessened.

**Figure 2.**
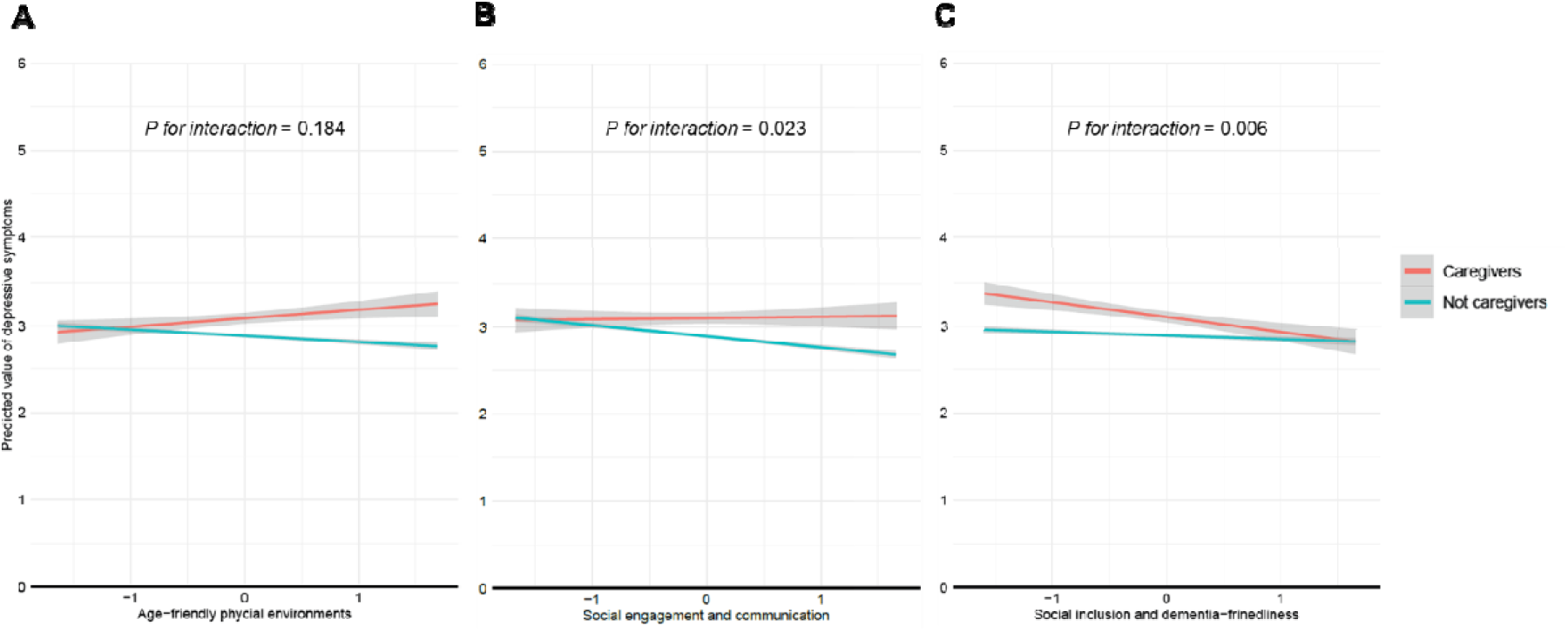
AFC scale scores and depressive symptoms according to caregiving status. AFC, age-friendly communities. The vertical axis indicates predicted value of depressive symptoms (GDS-15 score); the gray areas represent 95% confidence intervals. The scores of the AFC scale are shown in units per standard deviation (panels A, age-friendly physical environments; B, social engagement and communication; C, social inclusion and dementia-friendliness).

Supplementary Table 2 shows the subgroup analysis by gender on the association between caregiving status and depressive symptoms. Among both men and women, caregivers exhibited higher levels of depressive symptoms (Model 2). For men, the social inclusion and dementia-friendliness moderated the depressive symptoms of caregiving (Model 5), whereas for women, the moderation by the domain was not found (Model 5).

Supplementary Table 3 indicates the association between caregiving for persons with dementia and depressive symptoms. Caregivers of persons with dementia were more likely to have higher levels of depressive symptoms (Model 2), but the social inclusion and dementia-friendliness component moderated this association (Model 5). Although the social engagement and communication component tended to strengthen this association, no statistical evidence was found (Model 4).

Supplementary Table 4 shows the association between caregiving role and depressive symptoms. Primary caregivers were more likely to be depressed (Model 2). However, regarding the interaction between the caregiving role and AFC, although social inclusion and dementia-friendliness domain moderated the association between secondary caregivers and depressive symptoms, no statistical evidence was found to moderate the association between primary caregivers and depressive symptoms for any AFC domains (Models 3 to 5).

## Discussion and Implications

This study indicated that informal caregivers exhibited higher levels of depressive symptoms, and as hypothesized, this association was moderated by living in communities with socially inclusive and dementia-friendly elements of the AFC. Conversely, in communities with higher elements of physical environments or social engagement communication, the alleviation of caregivers’ depressive symptoms was not observed, which did not support our hypothesis. These findings provide insights into the roles of AFC for informal caregivers, suggesting the necessity of promoting the development and evaluation of AFC while considering the preservation of caregivers’ health and well-being.

The AFC reflects the commitment to a comprehensive and accessible community environment for the entire residents, and its inclusive and supportive features may provide resources to buffer caregiver stress. The WHO suggests that AFC can be beneficial for informal caregivers (WHO, 2007, 2015). The present study indicated that in the communities with age-friendliness, particularly elements of social inclusion and dementia-friendliness, informal caregivers’ depressive symptoms were moderated, suggesting the contextual effects of inclusiveness and dementia-friendliness toward caregivers’ mental health programs as well as other older residents. These findings add empirical evidence about the impact of AFC on caregivers and support the theoretical models of caregiving in terms of the buffering resources of age-friendly environments. Previous studies have suggested that caregivers’ mental health and well-being can be influenced by the community and neighborhood characteristics surrounding them, such as trust and connectedness (Mohanty et al., 2020; Noguchi et al., 2020a). These inclusive and supportive communities may alleviate caregiver-related stigma, such as a lack of understanding or contempt for the caregivers by society or the caregivers’ internalized shame or guilt (Zwar et al., 2025), encouraging caregivers to help-seeking from others, including medical and public sectors as well as neighbors and friends. These processes can lead to helping caregivers access social and emotional support. While these speculations lead to plausibility that the inclusive AFC elements are more specifically beneficial to caregivers, a socially vulnerable population, further investigations are required to elucidate the mechanisms. Still, our findings suggest the importance of promoting the AFC development with considering the inclusion and respect of informal caregivers of older adults and persons with dementia.

In the gender-based subgroup analysis, the domain of social inclusion and dementia-friendliness was more protective for male caregivers, but this was no longer observed in women. This difference may reflect the nature of gender differences in family caregiving. In general, men may be less prepared for caregiving than women due to a lack of care skills and knowledge (WHO et al, 2023). Therefore, male caregivers tend to actively outsource care and seek outside help (Gallicchio et al., 2002). This means that male caregivers may be more susceptible to environmental factors, and they might benefit more if the community social environment is inclusive and supportive for caregivers. Alternatively, traditional gender roles may relate. In Japan, the traditional belief that caregiving is the role of women still persists, and male caregivers are a minority, more likely to feel lonely and be socially vulnerable (Saito, 2017). Inclusive and supportive communities might have been beneficial to mental health by alleviating marginalization as a minority. Recently, informal caregivers have gradually diversified, including men, but internationally, family caregiving is still mainly performed out by women, who play a major role (WHO et al., 2023). The future requires exploring AFC that benefits all caregivers in common.

Our additional analysis indicated that the domain of social inclusion and dementia-friendliness moderated depressive symptoms of caregivers for persons with dementia. This domain also includes the elements of inclusive and positive attitudes toward people with dementia by the residents in the community. In such communities, there is less public stigma against dementia (Alzheimer’s Disease International, 2019), and caregivers for families with dementia may be less exposed to discrimination and stigma related to dementia, which may support their mental health. These results underscore the importance of promoting AFC considering dementia-friendly aspects and overcoming dementia stigma. Meanwhile, from the perspective of the caregiver’s role, the domain of social inclusion and dementia-friendliness was not necessarily mitigative against depressive symptoms. The results showed that depressive symptoms were alleviated for secondary caregivers, but it was not found for primary caregivers. Primary caregivers are more burdened and have severe mental health problems (Gorostiaga et al., 2022). The contextual influence of the community environments, alone, may not be sufficient to have a moderating effect on such severely burdened caregivers, and more substantial support may be necessary together, including the provision of medical and long-term care resources and services.

This study did not observe the moderating effects of caregivers’ depressive symptoms by communities with higher levels of physical environments and social engagement and communication domains. This means that such communities may not necessarily provide benefits to caregivers’ mental health specifically. The results contradict our hypothesis that AFC would also be caregiver-friendly, which suggests the possibility that the influence may vary depending on the AFC components. The domain of age-friendly physical environments covers accessibility to barrier-free buildings and transport resources. Certainly, physical barrier-free outdoor environments and accessibility to transport resources might help older adults with functional limitations and their caregivers daily living. However, evidence on the effects of physical aspects of the neighborhood environments on caregivers’ mental health is complex and limited. The literature suggests that the enrichment of the social environments is more important than the physical environments for caregivers’ mental health (Meyer et al., 2022). Additionally, caregivers also tend to demand social environments such as cohesion and support rather than physical environments as age-friendly aspects of the communities (Black et al., 2016); it suggests that the physical elements of the community environments might have limited benefits on caregivers’ psychological health.

Meanwhile, the domain of social engagement and communication reflects the level of activeness of older adults’ community group activities and civic engagement and people’s communication in the area. Several previous studies have suggested that higher levels of community-level social participation have a beneficial role in older people’s health and well-being (Haseda et al., 2018; Takeda et al., 2024). However, this positive influence has also been shown to be rather detrimental to some populations, such as those lacking social participation and those isolated (Amemiya et al., 2019). Communities with higher levels of social participation may accelerate the favorable impacts of health and well-being for active and healthy older adults, but those who tend to be isolated or have barriers to participation may have difficulty accessing the benefits. While caregivers’ social connections are powerful in protecting their mental health (Noguchi et al., 2020b; Oshio & Kan, 2016; Papastavrou et al., 2015), they also face a tendency to become isolated due to a number of factors, including time constraints, financial difficulties, and emotional toll (Hajek et al., 2021; Vasileiou et al., 2017; Velloze et al., 2022). In communities where healthy older adults are active in social participation, caregivers and their care recipients, socially vulnerable groups, may have barriers to benefit and may be, on the contrary, affected by feelings of exclusion and loneliness. Alternatively, because the AFC scale in the present study did not necessarily include participation in groups and gatherings for informal caregivers, it may not have successfully identified the positive effects of the social engagement and communication in the community on them. Regardless, these findings highlight the potential that AFC and its scale require updates considering caregiver-friendly. Further investigations are needed to identify environmental features appropriate for caregivers and evaluate their impact. Additionally, the present study only examined the influences of caregivers on one aspect of depressive symptoms, and thus, it is necessary to explore the multidimensional aspects of caregivers’ health, well-being, burdens, and social consequences.

This study has several limitations. First, the nature of the cross-sectional analysis could not address the causality. Further investigations using longitudinal data are required. Second, the items related to caregiving were based on self-reported information, such as the care recipient’s dementia diagnosis, which can result in misclassification. Third, this study had limited information on care recipients, with a lack of such data on their illness and functional levels. To test the influence of AFC on caregivers according to the care recipient status, further studies using dyadic data of the caregiver and the care recipient are needed. Fourth, the age of the informal caregivers was higher than that of the general population due to the data for those aged 65 and older. Thus, the results may not be applicable to younger caregivers. Still, given that older adults’ daily activities are more localized in their neighborhoods, this population may be suitable for understanding the influences of community environments. Finally, this study was based on data from a wide area in Japan, from urban to rural areas, but only 145 communities were ultimately analyzed to ensure the precision of the measurement of community variables, which may limit the representativeness of the entire area in Japan. Future investigations using nationwide community sample data are required to test the generalizability of the results.

## Conclusions

This study suggests that AFC aspects such as community social inclusion and dementia-friendliness have the potential to moderate the adverse association between informal caregiving and depressive symptoms. However, the aspects such as physical environments and social engagement and communication in the community may not necessarily be beneficial to caregivers’ mental health. These findings highlight the importance of promoting the AFC with inclusive and supportive perspectives for informal caregivers.

## Acknowledgments

We wish to express our sincere gratitude to the staff in the surveyed municipalities for their contributions. We also thank the members of the 300 BM and AFC working group of the Japan Gerontological Evaluation Study.

## Author contributions

TN conceptualized and designed the study, analyzed the data, interpreted the results, and drafted and revised the manuscript. SF helped to interpreted the results and reviewed and critically revised the manuscript. KI, SJ, TS reviewed and critically revised the manuscript. KK, the principal investigator of the JAGES, participated in data collection and study design and reviewed and critically revised the manuscript. TO participated in data collection and reviewed and critically revised the manuscript.

## Data availability

For the JAGES, all inquiries are to be addressed to the data management committee via. All JAGES datasets have ethical or legal restrictions for public deposition because of the inclusion of sensitive information about the human participants.

