## Supplemental Tables for "Role of age-friendly communities on the association between informal caregiving and depressive symptoms: a multilevel analysis using the Japan Gerontological Evaluation Study"

**Supplementary Table 1. AFC scale items**

| AFC domains | Items | Questions and responses |
| --- | --- | --- |
| **Age-friendly physical environments** |  |  |
| Outdoor spaces and buildings | Accessibility of barrier-free public spaces and buildings | Do you have any public facilities available for individuals experiencing difficulty walking or visual/hearing disabilities in your area? (responses: “a lot,” “somewhat,” “not much,” “not at all,” or “unknown”; 1 = “a lot” or “somewhat,” 0 = others) |
| Transportation | Accessibility of barrier-free streets | Do you have any streets available for individuals with wheelchairs, canes, or walkers to walk on in your neighborhood? (responses: “a lot,” “somewhat,” “not much,” “not at all,” or “unknown”; 1 = “a lot” or “somewhat,” 0 = others) |
| Outdoor spaces and buildings | Outdoor space suitable for exercise | Do you have any parks or sidewalks suitable for exercise or walking in your area? (responses: “a lot,” “somewhat,” “not much,” “not at all,” or “unknown"; 1 = "a lot" or "somewhat," 0 = others) |
| Transportation | Accessibility of barrier-free public transportation vehicles | Do you have any trains or buses available for individuals experiencing difficulty walking or visual/hearing disabilities in your area? (responses: “a lot,” “somewhat,” “not much,” “not at all,” or “unknown”; 1 = “a lot” or “somewhat,” 0 = others) |
| Transportation | Accessibility of public transportation stops | Do you have any train or subway stations in your neighborhood? (responses: “a lot,” “somewhat,” “not much,” “not at all,” or “unknown”; 1 = “a lot” or “somewhat,” 0 = others) |
| **Social engagement and communication** |  |  |
| Social participation | Participation in hobby groups | How often do you participate in hobby groups? (responses: “4 times or more a week,” “twice to three times a week,” “once a week,” “once to three times a month,” “sometimes a year,” or “none”; 1 = once or more a month, 0 = others) |
| Social participation | Participation in sports group and club | How often do you participate in sports group and club? (responses: “4 times or more a week,” “twice to three times a week,” “once a week,” “once to three times a month,” “sometimes a year,” or “none”; 1 = once or more a month, 0 = others) |
| Social participation | Participation in learning and cultural activities | How often do you participate in learning and cultural activities? (responses: “4 times or more a week,” “twice to three times a week,” “once a week,” “once to three times a month,” “sometimes a year,” or “none”: 1 = once or more a month, 0 = others) |
| Civic participation and employment | Participation in volunteer groups | How often do you participate in volunteer groups? (responses: “4 times or more a week,” “twice to three times a week,” “once a week,” “once to three times a month,” “sometimes a year,” or “none”; 1 = once or more a month, 0 = others) |
| Communication and information | Internet use | Have you used the Internet or e-mail in the past year? (responses: “none,” “sometimes a month,” “twice to three times a week,” or “almost everyday”; 1 = “sometimes a month” to “almost everyday,” 0 = “none”) |
| **Social inclusion and dementia-friendliness** | |  |
| Respect and social inclusion | Sence of belonging | Do you think you're respected by your neighbors and that you're a member of your community? (responses: “agree,” “somewhat agree,” “either,” “somewhat disagree,” or “disagree”; 1 = “agree” or “somewhat agree” or 0 = others) |
| Respect and social inclusion | Participation in community decisions | Do you participate in decision-making in your community by attending your neighborhood association or similar community meetings? (responses: “agree,” “somewhat agree,” “either,” “somewhat disagree,” or “disagree”; 1 = “agree” or “somewhat agree,” 0 = others) |
| Respect and social inclusion | Perception of community reciprocity | Do you think individuals living in your area try to help others in the most of situations? (responses: “agree,” “somewhat agree,” “either,” “somewhat disagree,” or “disagree”; 1 = “agree” or “somewhat agree,” 0 = others) |
| Communication and information | Frequency of meeting with friends | How often do you meet friends? (responses: “almost everyday,” “twice to three times a week,” “once a week,” “once to three times a month,” sometimes a year,” “none”; 1 = “once or more a month,” 0 = others) |
| Community support and health services | Community health care service | Do the local government offices and private companies in your area by and large offer welfare services necessary for your daily life and health? (responses: “agree,” “somewhat agree,” “either,” “somewhat disagree,” or “disagree”; 1 = “agree” or “somewhat agree,” 0 = others) |
| Dementia-friendliness | Social participation of people with dementia | Do you think individuals with dementia should take part in community activities and have some role in those activities? (responses: “agree,” “somewhat agree,” “either,” “somewhat disagree,” or “disagree”; 1 = “agree” or “somewhat agree,” 0 = others) |
| Dementia-friendliness | Support for families of people with dementia | If one of your family members were affected by dementia, would you like your neighbors and acquaintances to know about it so that you could get assistance from them? (responses: “agree,” “somewhat agree,” “either,” “somewhat disagree,” or “disagree”; 1 = “agree” or “somewhat agree,” 0 = others) |

*Note*: AFC, age-friendly community.

**Supplementary Table 2. A subgroup analysis by gender on association between caregiving status and depressive symptoms, based on multilevel linear regression analysis**

|  | Model 1 | |  | | Model 2 | | |  | | Model 3 | | |  | | Model 4 | | |  | | Model 5 | | |
| --- | --- | --- | --- | --- | --- | --- | --- | --- | --- | --- | --- | --- | --- | --- | --- | --- | --- | --- | --- | --- | --- | --- |
|  | Coef. (95% CI) | *P*-value | |  | | Coef. (95% CI) | *P*-value | |  | | Coef. (95% CI) | *P*-value | |  | | Coef. (95% CI) | *P*-value | |  | | Coef. (95% CI) | *P*-value |
| ***Men (n = 4,706)*** |  |  | |  | |  |  | |  | |  |  | |  | |  |  | |  | |  |  |
| ***Fixed effects*** |  |  | |  | |  |  | |  | |  |  | |  | |  |  | |  | |  |  |
| **Caregiving status** |  |  | |  | |  |  | |  | |  |  | |  | |  |  | |  | |  |  |
| Not caregivers | Reference |  | |  | | Reference |  | |  | | Reference |  | |  | | Reference |  | |  | | Reference |  |
| Caregivers | 0.58 (0.28, 0.88) | < 0.001 | |  | | 0.56 (0.28, 0.85) | < 0.001 | |  | | 0.56 (0.27, 0.85) | < 0.001 | |  | | 0.57 (0.28, 0.86) | < 0.001 | |  | | 0.56 (0.27, 0.84) | < 0.001 |
| **AFC scale** |  |  | |  | |  |  | |  | |  |  | |  | |  |  | |  | |  |  |
| Age-friendly physical environments |  |  | |  | | 0.05 (-0.12, 0.23) | 0.541 | |  | | 0.03 (-0.15, 0.21) | 0.747 | |  | | 0.06 (-0.12, 0.23) | 0.521 | |  | | 0.05 (-0.12, 0.22) | 0.561 |
| Social engagement and communication |  |  | |  | | -0.20 (-0.56, 0.15) | 0.268 | |  | | -0.19 (-0.55, 0.16) | 0.282 | |  | | -0.24 (-0.6, 0.13) | 0.201 | |  | | -0.20 (-0.55, 0.16) | 0.278 |
| Social inclusion and dementia-friendliness |  |  | |  | | -0.31 (-0.54, -0.07) | 0.011 | |  | | -0.30 (-0.54, -0.07) | 0.011 | |  | | -0.31 (-0.54, -0.07) | 0.011 | |  | | -0.25 (-0.49, -0.01) | 0.041 |
| **Caregiving status x AFC scale** |  |  | |  | |  |  | |  | |  |  | |  | |  |  | |  | |  |  |
| Caregiver x Age-friendly physical environments |  |  | |  | |  |  | |  | | 0.25 (-0.18, 0.68) | 0.259 | |  | |  |  | |  | |  |  |
| Caregivers x Social engagement and communication |  |  | |  | |  |  | |  | |  |  | |  | | 0.41 (-0.34, 1.16) | 0.282 | |  | |  |  |
| Caregivers x Social inclusion and dementia-friendliness |  |  | |  | |  |  | |  | |  |  | |  | |  |  | |  | | -0.63 (-1.16, -0.11) | 0.018 |
| ***Random effects*** |  |  | |  | |  |  | |  | |  |  | |  | |  |  | |  | |  |  |
| Community-level variance | 0.1315 |  | |  | | 0.07277 |  | |  | | 0.07269 |  | |  | | 0.07318 |  | |  | | 0.07108 |  |
| ***Women (n = 5,609)*** |  |  | |  | |  |  | |  | |  |  | |  | |  |  | |  | |  |  |
| ***Fixed effects*** |  |  | |  | |  |  | |  | |  |  | |  | |  |  | |  | |  |  |
| **Caregiving status** |  |  | |  | |  |  | |  | |  |  | |  | |  |  | |  | |  |  |
| Not caregivers | Reference |  | |  | | Reference |  | |  | | Reference |  | |  | | Reference |  | |  | | Reference |  |
| Caregivers | 0.40 (0.12, 0.68) | 0.005 | |  | | 0.44 (0.18, 0.71) | 0.001 | |  | | 0.45 (0.18, 0.72) | 0.001 | |  | | 0.45 (0.19, 0.72) | < 0.001 | |  | | 0.44 (0.17, 0.71) | 0.001 |
| **AFC scale** |  |  | |  | |  |  | |  | |  |  | |  | |  |  | |  | |  |  |
| Age-friendly physical environments |  |  | |  | | 0.01 (-0.15, 0.17) | 0.904 | |  | | -0.02 (-0.18, 0.15) | 0.849 | |  | | 0.01 (-0.15, 0.17) | 0.902 | |  | | 0.01 (-0.15, 0.17) | 0.904 |
| Social engagement and communication |  |  | |  | | -0.03 (-0.36, 0.30) | 0.863 | |  | | -0.04 (-0.37, 0.29) | 0.830 | |  | | -0.08 (-0.42, 0.26) | 0.659 | |  | | -0.03 (-0.36, 0.30) | 0.865 |
| Social inclusion and dementia-friendliness |  |  | |  | | -0.21 (-0.43, 0.00) | 0.055 | |  | | -0.21 (-0.43, 0.01) | 0.059 | |  | | -0.21 (-0.43, 0.01) | 0.057 | |  | | -0.22 (-0.44, 0.01) | 0.058 |
| **Caregiving status x AFC scale** |  |  | |  | |  |  | |  | |  |  | |  | |  |  | |  | |  |  |
| Caregivers x Age-friendly physical environments |  |  | |  | |  |  | |  | | 0.27 (-0.14, 0.69) | 0.197 | |  | |  |  | |  | |  |  |
| Caregivers x Social engagement and communication |  |  | |  | |  |  | |  | |  |  | |  | | 0.43 (-0.25, 1.11) | 0.216 | |  | |  |  |
| Caregivers x Social inclusion and dementia-friendliness |  |  | |  | |  |  | |  | |  |  | |  | |  |  | |  | | 0.03 (-0.47, 0.52) | 0.914 |
| ***Random effects*** |  |  | |  | |  |  | |  | |  |  | |  | |  |  | |  | |  |  |
| Community-level variance | 0.04637 |  | |  | | 0.002731 |  | |  | | 0.001432 |  | |  | | 0.001853 |  | |  | | 0.002744 |  |

*Note*: AFC, age-friendly community; Coef., unstandardized regression coefficient; CI, confidence interval.

Model 1: crude model; Model 2: adjusted by age, living arrangement, marital status, equivalent household income, home ownership, household total assets, work frequency, number of illnesses, basic and instrumental activities of daily living, population density, aging proportion, low education proportion, care resource, and AFC scale score; Model 3 to 5: added the interaction terms to Model 2 (Model 3: caregiving status x Age-friendly physical environments; Model 4: caregiving status x Social engagement and communication; Model 4: caregiving status x Social inclusion and dementia-friendliness).

Random effects for the null model (community-level variance) = 0.09613 for men and 0.09330 for women.

The AFC scale scores were estimated per ten percent points.

Missing data were imputed by random forest imputation algorithm.

**Supplementary Table 3. Association between caregiving status (for person with dementia) and depressive symptoms, based on multilevel linear regression analysis**

|  | Model 1 | |  | Model 2 | |  | Model 3 | |  | Model 4 | |  | Model 5 | |
| --- | --- | --- | --- | --- | --- | --- | --- | --- | --- | --- | --- | --- | --- | --- |
|  | Coef. (95% CI) | *P*-value |  | Coef. (95% CI) | *P*-value |  | Coef. (95% CI) | *P*-value |  | Coef. (95% CI) | *P*-value |  | Coef. (95% CI) | *P*-value |
| ***Fixed effects*** |  |  |  |  |  |  |  |  |  |  |  |  |  |  |
| **Caregiving status** |  |  |  |  |  |  |  |  |  |  |  |  |  |  |
| Not caregivers | Reference |  |  | Reference |  |  | Reference |  |  | Reference |  |  | Reference |  |
| Caregivers for person without dementia | 0.19 (-0.08, 0.45) | 0.169 |  | 0.16 (-0.10, 0.42) | 0.225 |  | 0.17 (-0.09, 0.42) | 0.205 |  | 0.16 (-0.1, 0.42) | 0.223 |  | 0.16 (-0.1, 0.42) | 0.219 |
| Caregivers for person with dementia | 0.33 (0.07, 0.58) | 0.012 |  | 0.33 (0.08, 0.58) | 0.009 |  | 0.33 (0.08, 0.58) | 0.009 |  | 0.33 (0.08, 0.58) | 0.009 |  | 0.31 (0.06, 0.56) | 0.016 |
| **AFC scale** |  |  |  |  |  |  |  |  |  |  |  |  |  |  |
| Age-friendly physical environments |  |  |  | 0.00 (-0.11, 0.11) | 0.998 |  | -0.01 (-0.13, 0.10) | 0.830 |  | 0.00 (-0.11, 0.11) | 0.966 |  | 0.00 (-0.11, 0.11) | 0.984 |
| Social engagement and communication |  |  |  | -0.05 (-0.27, 0.17) | 0.659 |  | -0.05 (-0.28, 0.17) | 0.655 |  | -0.11 (-0.34, 0.12) | 0.357 |  | -0.05 (-0.27, 0.18) | 0.672 |
| Social inclusion and dementia-friendliness |  |  |  | -0.23 (-0.38, -0.08) | 0.003 |  | -0.23 (-0.38, -0.07) | 0.003 |  | -0.23 (-0.38, -0.07) | 0.003 |  | -0.17 (-0.33, -0.02) | 0.031 |
| **Caregiving status x AFC scale** |  |  |  |  |  |  |  |  |  |  |  |  |  |  |
| Caregivers for person without dementia x Age-friendly physical environments |  |  |  |  |  |  | 0.22 (-0.18, 0.63) | 0.279 |  |  |  |  |  |  |
| Caregivers for person with dementia x Age-friendly physical environments |  |  |  |  |  |  | 0.02 (-0.35, 0.40) | 0.900 |  |  |  |  |  |  |
| Caregivers for person without dementia x Social engagement and communication |  |  |  |  |  |  |  |  |  | 0.54 (-0.15, 1.23) | 0.126 |  |  |  |
| Caregivers for person with dementia x Social engagement and communication |  |  |  |  |  |  |  |  |  | 0.51 (-0.14, 1.15) | 0.125 |  |  |  |
| Caregivers for person without dementia x Social inclusion and dementia-friendliness |  |  |  |  |  |  |  |  |  |  |  |  | -0.48 (-0.95, -0.01) | 0.047 |
| Caregivers for person with dementia x Social inclusion and dementia-friendliness |  |  |  |  |  |  |  |  |  |  |  |  | -0.61 (-1.09, -0.13) | 0.012 |
| ***Random effects*** |  |  |  |  |  |  |  |  |  |  |  |  |  |  |
| Community-level variance | 0.08705 |  |  | 0.04259 |  |  | 0.04246 |  |  | 0.04339 |  |  | 0.04373 |  |

*Note*: AFC, age-friendly community; Coef., unstandardized regression coefficient; CI, confidence interval.

Model 1: crude model; Model 2: adjusted by age, gender, living arrangement, marital status, equivalent household income, home ownership, household total assets, work frequency, number of illnesses, basic and instrumental activities of daily living, population density, aging proportion, low education proportion, care resource, and AFC scale score; Model 3 to 5: added the interaction terms to Model 2 (Model 3: caregiving status x Age-friendly physical environments; Model 4: caregiving status x Social engagement and communication; Model 4: caregiving status x Social inclusion and dementia-friendliness).

Random effects for the null model (community-level variance) = 0.08898.

The AFC scale scores were estimated per ten percent points.

Missing data were imputed by random forest imputation algorithm.

**Supplementary Table 4. Association between caregiving status (role) and depressive symptoms, based on multilevel linear regression analysis**

|  | Model 1 | |  | Model 2 | |  | Model 3 | |  | Model 4 | |  | Model 5 | |
| --- | --- | --- | --- | --- | --- | --- | --- | --- | --- | --- | --- | --- | --- | --- |
|  | Coef. (95% CI) | *P*-value |  | Coef. (95% CI) | *P*-value |  | Coef. (95% CI) | *P*-value |  | Coef. (95% CI) | *P*-value |  | Coef. (95% CI) | *P*-value |
| ***Fixed effects*** |  |  |  |  |  |  |  |  |  |  |  |  |  |  |
| **Caregiving status** |  |  |  |  |  |  |  |  |  |  |  |  |  |  |
| Not caregiver | Reference |  |  | Reference |  |  | Reference |  |  | Reference |  |  | Reference |  |
| Secondary caregiver | 0.08 (-0.14, 0.30) | 0.482 |  | 0.04 (-0.17, 0.26) | 0.701 |  | 0.05 (-0.17, 0.26) | 0.674 |  | 0.05 (-0.16, 0.27) | 0.630 |  | 0.04 (-0.18, 0.25) | 0.719 |
| Primary caregiver | 0.57 (0.28, 0.86) | < 0.001 |  | 0.62 (0.34, 0.90) | < 0.001 |  | 0.62 (0.34, 0.90) | < 0.001 |  | 0.62 (0.34, 0.90) | < 0.001 |  | 0.62 (0.34, 0.90) | < 0.001 |
| **AFC scale** |  |  |  |  |  |  |  |  |  |  |  |  |  |  |
| Age-friendly physical environments |  |  |  | 0.00 (-0.11, 0.11) | 0.975 |  | 0.00 (-0.12, 0.11) | 0.972 |  | 0.00 (-0.11, 0.11) | 0.971 |  | 0.00 (-0.11, 0.11) | 0.975 |
| Social engagement and communication |  |  |  | -0.05 (-0.27, 0.17) | 0.659 |  | -0.05 (-0.27, 0.18) | 0.693 |  | -0.07 (-0.30, 0.16) | 0.561 |  | -0.05 (-0.27, 0.18) | 0.668 |
| Social inclusion and dementia-friendliness |  |  |  | -0.23 (-0.38, -0.08) | 0.003 |  | -0.23 (-0.38, -0.08) | 0.003 |  | -0.23 (-0.38, -0.08) | 0.003 |  | -0.19 (-0.35, -0.04) | 0.015 |
| **Caregiving status x AFC scale** |  |  |  |  |  |  |  |  |  |  |  |  |  |  |
| Secondary caregiver x Age-friendly physical environments |  |  |  |  |  |  | 0.25 (-0.08, 0.58) | 0.138 |  |  |  |  |  |  |
| Primary caregiver x Age-friendly physical environments |  |  |  |  |  |  | -0.39 (-0.84, 0.05) | 0.084 |  |  |  |  |  |  |
| Secondary caregiver x Social engagement and communication |  |  |  |  |  |  |  |  |  | 0.34 (-0.24, 0.91) | 0.251 |  |  |  |
| Primary caregiver x Social engagement and communication |  |  |  |  |  |  |  |  |  | -0.14 (-0.87, 0.59) | 0.698 |  |  |  |
| Secondary caregiver x Social inclusion and dementia-friendliness |  |  |  |  |  |  |  |  |  |  |  |  | -0.50 (-0.90, -0.09) | 0.016 |
| Primary caregiver x Social inclusion and dementia-friendliness |  |  |  |  |  |  |  |  |  |  |  |  | 0.03 (-0.50, 0.56) | 0.906 |
| ***Random effects*** |  |  |  |  |  |  |  |  |  |  |  |  |  |  |
| Community-level variance | 0.08853 |  |  | 0.04368 |  |  | 0.04334 |  |  | 0.0434 |  |  | 0.04413 |  |

*Note*: AFC, age-friendly community; Coef., unstandardized regression coefficient; CI, confidence interval.

Model 1: crude model; Model 2: adjusted by age, gender, living arrangement, marital status, equivalent household income, home ownership, household total assets, work frequency, number of illnesses, basic and instrumental activities of daily living, population density, aging proportion, low education proportion, care resource, and AFC scale score; Model 3 to 5: added the interaction terms to Model 2 (Model 3: caregiving status x Age-friendly physical environments; Model 4: caregiving status x Social engagement and communication; Model 4: caregiving status x Social inclusion and dementia-friendliness).

Random effects for the null model (community-level variance) = 0.08898.

The AFC scale scores were estimated per ten percent points.

Missing data were imputed by random forest imputation algorithm.
